# Convolutional neural network reveals frequency content of medio-lateral COM body sway to be highly predictive of Parkinson’s disease

**DOI:** 10.1101/2023.05.26.23289242

**Authors:** David Engel, R. Stefan Greulich, Alberto Parola, Kaleb Vinehout, Stefan Dowiasch, Josefine Waldthaler, Lars Timmermann, Constantin A. Rothkopf, Frank Bremmer

## Abstract

Postural instability as a symptom of progressing Parkinson’s disease (PD) greatly reduces quality of life. Hence, early detection of postural impairments is crucial to facilitate interventions. Our aim was to use a convolutional neural network (CNN) to differentiate people with early to mid-stage PD from healthy age-matched individuals based on spectrogram images obtained from their body movement. We hypothesized the time-frequency content of body sway to be predictive of PD, even when impairments are not yet manifested in day-to-day postural control. We tracked their center of pressure (COP) using a Wii Balance Board and their full-body motion using a Microsoft Kinect, out of which we calculated the trajectory of their center of mass (COM). We used 30 s-snippets of motion data from which we acquired wavelet-based time-frequency spectrograms that were fed into a custom-built CNN as labeled images. We used binary classification to have the network differentiate between individuals with PD and controls (n=15, respectively). Classification performance was best when the medio-lateral motion of the COM was considered. Here, our network reached an average predictive accuracy of 98.45 % with a receiver operating characteristic area under the curve of 1.0. Moreover, an explainable AI approach revealed high frequencies in the postural sway data to be most distinct between both groups. Our findings suggest a CNN classifier based on cost-effective and conveniently obtainable posturographic data to be a promising approach to detect postural impairments in early to mid-stage PD and to gain novel insight into the subtle characteristics of impairments at this stage of the disease.

## Introduction

With the ongoing demographic transition towards an increasingly elderly population in western industrialized countries, neurodegenerative diseases, typically occurring with old age, are becoming increasingly common. Second only to Alzheimer’s, Parkinson’s disease (PD) currently affects around 1% of the population over 60 (Elbaz et al., 2016; Tysnes & Storstein, 2017) and is predicted to affect around 3% of the world population over 65 by 2030 (Palakurthi & Burugupally, 2019). Postural instability is considered one of the most disabling features of PD and at later stages highly increases the risk of falls, thereby strongly reducing the quality of life of those affected (Benatru et al., 2008; Bloem, 1992; Doná et al., 2016; Grimbergen et al., 2009; Horak, 2006; Hwang et al., 2016; Kim et al., 2013). There is still no cure for the disease, which makes it important to detect and predict impending balance impairments as early as possible. This would allow for timely countermeasures to facilitate treatment and decelerate symptoms, as there is evidence for positive effects of exercise and specific balance training on postural control (Allen et al., 2022), and it has also been shown that neuroprotective and neuromodulatory therapies may have the potential to delay disease progression in the future (Holford & Nutt, 2008; Sarkar et al., 2016).

Most of the symptoms are to this date evaluated in simple gold-standard clinical tests, performed by a clinician with the help of reference frameworks like the Movement Disorder Society Unified Parkinson’s Disease Rating Scale (MDS-UPDRS) (Goetz et al., 2008; Landers et al., 2008; Siderowf et al., 2002) and the Hoehn and Yahr scale (Hoehn & Yahr, 1967). Even though scales like the UPDRS score have been attested a good test-retest reliability (Siderowf et al., 2002), most clinical balance ratings within these frameworks remain (semi-)subjective and might be biased by various outside circumstances that do not reflect the current state of the disease (Evers et al., 2019; Liu et al., 2022). For instance, common clinical balance tests have been shown to be insensitive to mild impairments (Ebersbach et al., 2006) and to have a poor sensitivity as well as trial-to-trial stability (Dibble & Lange, 2006; Luque-Casado et al., 2021). In addition, motor symptoms visible to a clinician during these tests typically occur at later stages of the disease, while it has been suggested that postural impairments might already be present in the early motor phase and that they might bear the potential as a pre-diagnostic tool to detect the disease (Beuter et al., 2008; Chastan et al., 2008). Hence, there is a strong need for objective and easy to obtain measures of postural control to assist clinical testing of PD (Palakurthi & Burugupally, 2019).

This is why in recent years many studies were trying to find objective static and dynamic measures of postural control that distinguish individuals with PD from healthy controls (HC). However, results thus far have been contradictory, which has mainly been attributed to the wide heterogeneity of affected individuals and wide variety of study designs (Kamieniarz et al., 2018). A further limitation in the case of static balance control remains that most studies so far only investigated spatiotemporal aspects of posturographic balance measures in clinical populations (Kamieniarz et al., 2018). Though, a few recent studies successfully distinguished between individuals with PD at various stages and HC as well as between PD subtypes based on the frequency content of their COP trajectories during quiet stance (Kamieniarz et al., 2021; Rezvanian et al., 2018; Rocchi et al., 2006). This makes evaluation of the frequency domain of body sway signals during static balance control a promising candidate to detect features of static posturography that are unique to PD.

There has been a recent rise in using machine learning techniques to evaluate various aspects of PD. In a recent review, Mei and colleagues surveyed 209 studies within the machine learning literature focusing on classification of PD. The most used data types were voice recordings, gait patterns, hand writings and MRI imaging, resulting in prediction accuracies between 80% and 100% (Mei et al., 2021). In terms of static balance assessments, different machine learning algorithms have been compared on various features of COP sway recorded during quiet standing, leading to prediction accuracies between 64% and 83.9% (Fadil et al., 2021; Li et al., 2020). Recent studies have also used deep learning approaches, where computational models are composed of multiple processing layers to learn representations of data with multiple levels of abstraction (Lecun et al., 2015). Convolutional neural networks (CNN) constitute a widely established deep learning algorithm for image classification, with an architecture somewhat similar to the ventral cortical stream of the human visual system and consist of various layers that use learnable filters for feature extraction (Krizhevsky et al., 2017; Lecun et al., 2015). CNN approaches have been used in the context of PD based on brain imaging data like SPECT, MRI, and PET, but also successfully on physiological data like gait patterns, handwritings, and speech (Loh, Hong, et al., 2021). CNNs also show great potential when being used on spectrogram data based on time-frequency representations of spatiotemporal signals. This has been applied to audio signals (Dayal et al., 2022), speech recognition (Badshah et al., 2017), but even in the context of gait classification (Jung et al., 2019). In clinical contexts with PD, recent studies have used this approach on EEG data (Khare et al., 2021; Loh, Ooi, et al., 2021).

Taken together, these studies prove a wide range of potential applications of machine learning to determine biomarkers of PD from diagnostic data. However, the majority utilized data obtained in laboratory experiments which require expensive hardware and trained experts performing elaborate protocols, resources most often not available to general practitioners or medical practitioners, especially in disenfranchised countries (Dotchin & Walker, 2012). Hence, there remains demand for a high-performing approach which is based on easily and quickly obtainable data that does not require elaborate experimental setups. Given the persisting lack of suitable objective measures to assist clinical balance assessment in PD and the promising application of machine learning methods to various aspects of the disease, the aim of our study was to develop a CNN classifier trained on short excerpts of static posturographic data that will be able to generalize for new subjects based on a quick and easy assessment. For this purpose, we recorded the COP and COM trajectories of individuals with PD and age-matched HC during quiet standing using mobile and cost-effective devices (Engel, Schwenk, et al., 2021; Engel, Student, et al., 2021; Student et al., 2022) and trained a custom-built CNN to distinguish both groups based on the time-frequency content represented as spectrogram images. Moreover, along the urgent need of explainable AI, we used a GradCAM approach (Selvaraju et al., 2020) to understand which frequencies in the postural sway data were vital to the classifier’s decision. We hypothesized that a CNN trained on time-frequency spectrogram images acquired during short episodes of quiet stance can reliably distinguish individuals with early to mid-stage PD from age-matched HC subjects and thus bears great potential not only for future clinical applications, but also for a better understanding of standing posture in PD.

## Results

### Prediction Performance

Table 1 displays the prediction performance of our network for the COP and COM measures in the anterior-posterior (a-p) and medio-lateral (m-l) directions, respectively, averaged over 250 training sessions with randomized train- and test set assignments of individual subjects (see Methods). While the network was able to achieve high accuracies in single training sessions when trained on the COP data, average test accuracy reached about 75 % for the m-l direction and only around 70 % for the a-p direction, with a large heterogeneity in performance showing in the high standard deviations (SD) of 11.3 % and 14.3 %, respectively. The receiver operating characteristics area under the curves (ROC AUC) reached values of 0.8 and 0.75, respectively, again with large variability (Table 1). The network performed significantly better when trained on the COM data. Here, average test accuracy was 90.2 % for data obtained from the a-p direction, however, again with a high SD of 10.2 %, indicating large variability in performance. In this direction, ROC AUC was 0.95 with a SD of 0.09. When trained on the COM data obtained from the m-l direction, the network’s performance was by far best, reaching an average test accuracy of 98.5 % with a small SD of 3.6 %, indicating an excellent predictive performance which proved to be consistent across training sessions. Here, the ROC AUC was 0.995 on average, with a small SD of 0.026, again indicating highly reliable performance. Figure 1 shows our network’s performance based on the medio-lateral COM data during training. Evaluation on both the training and test sets shows a monotonous decrease of the loss function after about 200 training epochs. For the test set, containing images that the network has never seen before, this indicates robust training without overfitting on the training data. On average, training and test accuracies start increasing after about 50-100 training epochs, both reaching a plateau of maximum performance after about 600 epochs (Fig. 1A). Remarkably, our network’s performance was almost identical between the train and the test set. This indicates that it performs as well on novel data as it does on data it has been trained with. Panel B of Figure 1 shows the receiver operating characteristic curves of the five best-performing models. These revealed that our network can reach excellent sensitivity for all thresholds of false negative rates, indicating close to perfect skill in the decision process.

**Table 1:** CNN performance on data from each postural parameter and corresponding direction. Data based on 250 models, each trained with data from different subjects in the train and test sets.

|  | Test Accuracy<br>Average + SD | Test Accuracy<br>Best | Test Accuracy<br>Worst | ROC AUC<br>Average + SD | ROC AUC<br>Best | ROC AUC<br>Worst |
| --- | --- | --- | --- | --- | --- | --- |
| **COP<sub>ml</sub>** | 74.57 +/- 11.30 % | 98.54 % | 53.75 % | 0.7947 +/-<br>0.1261 | 0.9995 | 0.5297 |
| **COP<sub>ap</sub>** | 70.25 +/- 14.25 % | 98.13 % | 46.46 % | 0.7466 +/-<br>0.1559 | 0.9995 | 0.4379 |
| **COM<sub>ml</sub>** | 98.45 +/- 3.61 % | 100 % | 62.45 % | 0.9954 +/-<br>0.0259 | 1 | 0.6777 |
| **COM<sub>ap</sub>** | 90.21 +/- 10.22 % | 100 % | 58.11 % | 0.9455 +/-<br>0.0865 | 1 | 0.6171 |

**Fig. 1:**
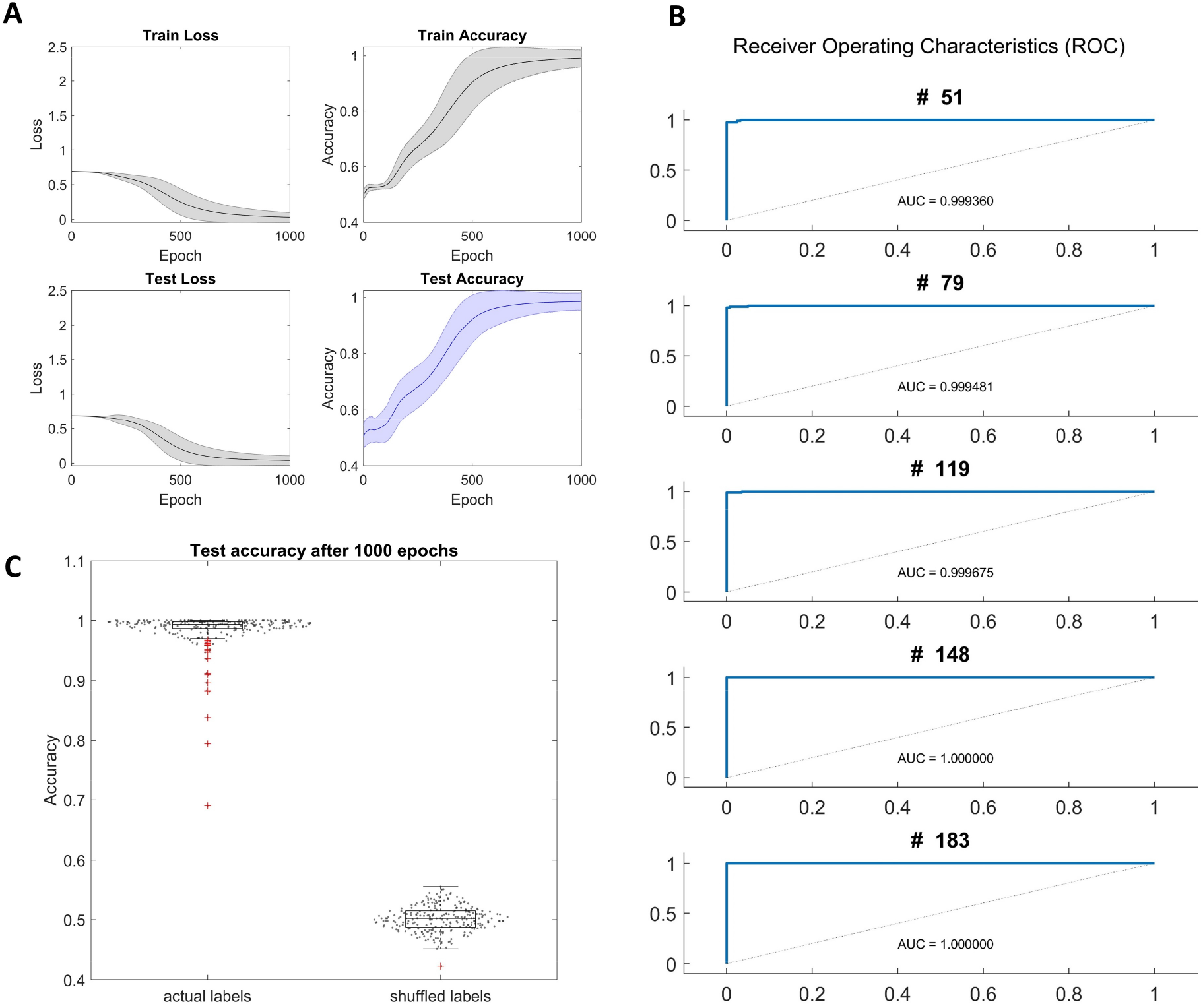
Network performance on medio-lateral COM data. Panel A: Network performance indicated by total loss and classification accuracy evaluated on the train and test sets over 1000 epochs of training. Solid lines represent the average across 250 models with different subjects assigned to the train and test sets in each case. Shaded areas indicate standard error. Panel B: ROC curves of the 5 best-performing models evaluated after 1000 epochs. Panel C: Test accuracy after 1000 epochs of training. Results from 250 models trained with correctly labeled data versus shuffled labels. Each model trained with different subjects in the train and test set. Grey dots indicate single samples. Red crosses indicate outliers.

To ascertain that our network’s performance was based on an actual difference in the data between both groups, we created additional data sets with shuffled labels on which we trained another 250 models, again each time with random assignment of participants to the train and test set. The performance of the models trained with randomly labeled data is compared to the performance when trained on actual data in Figure 1C. Without label information, test accuracy after 1000 epochs of training remained around chance level. There was a significant difference in test accuracy across 250 models between the actual and shuffled data sets (p < 0.0001).

### Gradient class activation maps (GradCAM)

Since our network reached excellent performance on the spectrograms obtained from medio-lateral sway of the COM, we applied gradient-based class activation maps (GradCAM) to obtain insight into the decision process of the network, i.e., to understand which sections of the spectrograms were relevant for the network’s decision whether data came from an individual with PD or a HC. We calculated the GradCAMs for the five best-performing models (test accuracies of 100%). For each model, we used the complete batch of test set images (between 402 and 474 depending on the model, due to some trials being discarded, see Methods) and averaged them over each decoded class. The results can be seen in Figure 2. As all models show, in cases where the image was classified to come from an individual with PD (Fig. 2, right column), most pixels representing the higher frequency bands between about 0.5 Hz and 5 Hz were given the largest weights. Three of the models (#51, #119, #148) had their largest weights around 1 Hz, whereas the remaining two (#79, #183) had the largest gradients at frequencies above 2 Hz. Noteworthy, weights remained consistent over time in all models. When the models classified a control subject (Fig. 2, left column), weights were on average much lower and more evenly distributed across the spectrograms. However, here, all models shared a narrow band of highest gradients at around 2.5 Hz.

**Fig. 2.**
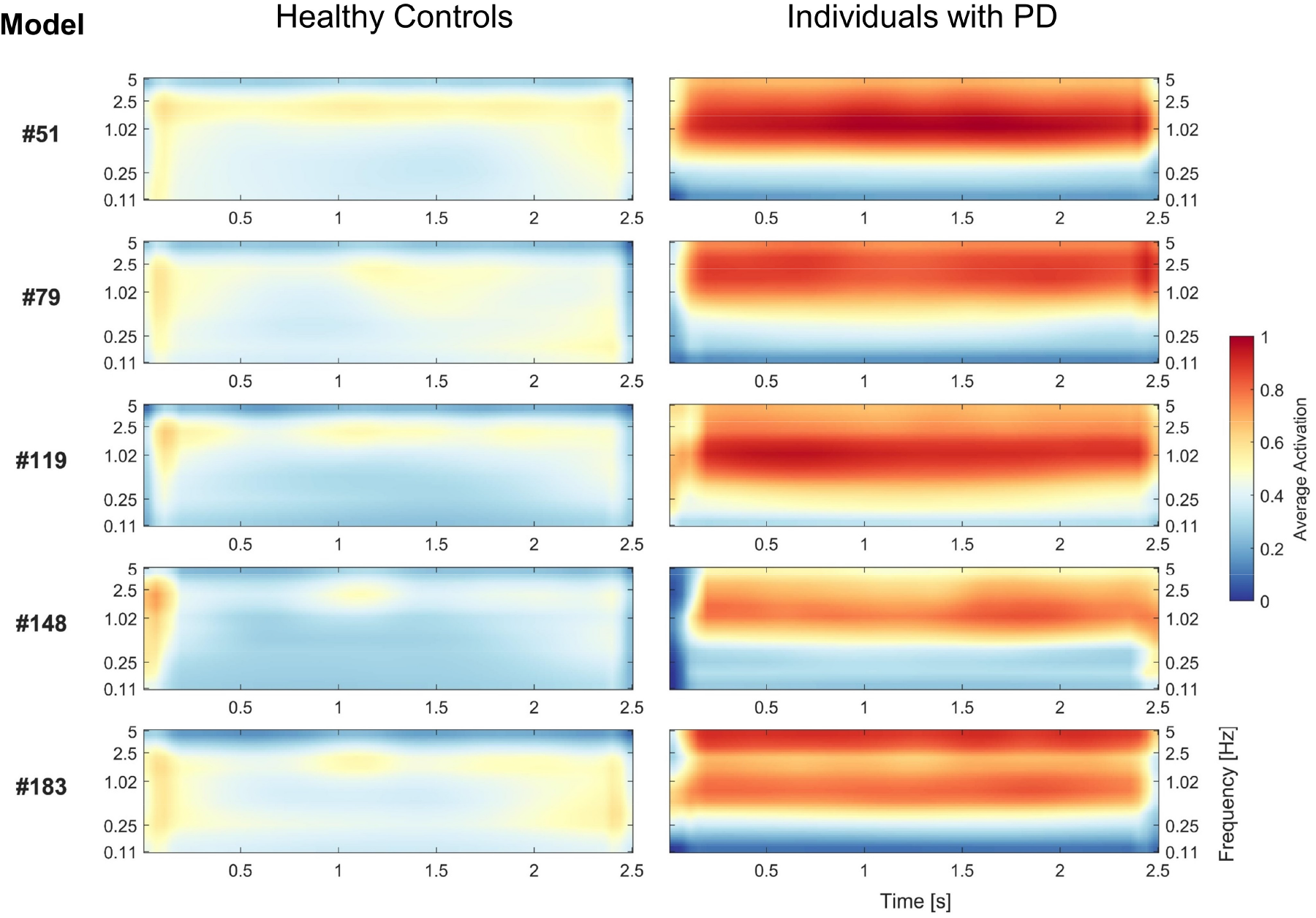
Gradient class activation maps (GradCAM) of the last convolutional layer in the five best-performing models trained on medio-lateral COM data. Left panels show average GradCAMs for HC, right panels show GradCAMs for individuals with PD. Consistent across models, when an individual with PD was classified, large weights were given to the higher frequencies between 0.5 Hz and 5 Hz. When a HC was classified, average weights were much lower and more evenly distributed across the spectrograms.

## Discussion

In our study, we developed a CNN classifier trained on short (5s) excerpts of static posturographic data obtained from individuals with PD and healthy age-matched control subjects with mobile and cost-effective equipment. We used a custom-built network to distinguish between individuals of both groups based on the time-frequency content of their COP and COM trajectories during quiet stance, represented as spectrogram images. Moreover, we used GradCAMs as an explainable AI approach to investigate which sections of the spectrograms were vital for each model’s decision. We found our network to reach excellent classification performance when trained on medio-lateral body sway data obtained from participants’ COM. Our best-performing models exhibited a test accuracy of 100 % with a ROC AUC of 1.0. This made us confirm our hypothesis that there are differences in the frequency content of body sway signals between both groups that are reliably detectable with a computer vision approach. Besides, applying the GradCAM technique allowed us to obtain interpretable insight into the network’s decision process by revealing that frequency bands between 0.5 Hz and 5 Hz were most predictive for individuals with PD.

We tested our network on postural sway data during quiet standing analyzed at two common postural sway parameters, the COP and COM. This revealed that decision performance was by far best when based on the COM data in the medio-lateral direction (Table 1). The fact that our network performed best on data obtained from medio-lateral body sway appears plausible as PD motor symptoms are lateralized, especially in early disease stages (Djaldetti et al., 2006; Riederer et al., 2018). This finding indicates a larger difference in the time-frequency spectrograms between the groups and confirms various studies in the literature that used conventional analyses in the spatiotemporal realm. For instance, it has been found that individuals with PD show increased postural sway and stochastic activity in their medio-lateral postural sway in static balance assessments (Błaszczyk et al., 2007; Błaszczyk & Orawiec, 2011; Chastan et al., 2008; Mitchell et al., 1995; Stylianou et al., 2011), which lead to the suggestion that lateral instability may be an important posturographic marker of functional balance impairment in PD (Błaszczyk et al., 2007; Mitchell et al., 1995; Rocchi et al., 2006). This has been confirmed in a recent study using spatiotemporal analyses, where it was found that investigation of the medio-lateral direction of postural sway was best suited to differentiate between individuals with PD and HC (Sebastia-Amat et al., 2023). Moreover, as all people with PD who participated in our study received L-dopa at the time of testing, there is evidence that L-dopa has negative effects on medio-lateral stability more so than on anterior-posterior stability, possibly increasing distinguishability for our deep learning models (e.g., Rocchi et al., 2006).

Our GradCAMs (Fig. 2) revealed that, on average, our models ‘looked’ at different areas in the spectrograms each time they classified a healthy subject, but that they ‘looked’ at very similar areas each time they classified a person with PD, which indicates that the latter share a common postural trait. This postural trait seems to be reflected in the higher frequency components of their postural sway during quiet stance. Biomechanically, higher frequencies have been associated with increased stiffness around the ankle joint (Warnica et al., 2014), that might be explained by the increased rigidity commonly associated with PD (Chastan et al., 2008; Engel, Student, et al., 2021). In terms of central-nervous processing of balance control, three frequency bands are usually associated with involvement of specific neuronal loops: 0 - 0.5 Hz for visuo-vestibular regulation, 0.5 - 2 Hz for cerebellar participation, and 2 - 20 Hz for proprioceptive participation (Fadil et al., 2021; Paillard & Noé, 2015). Since our models detected differences between individuals with PD and HC mostly in the frequency bands between 0.5 Hz and 5 Hz, this might reflect impairments in the latter two central-nervous loops: The lower range of the most distinctive frequency bands might represent altered cerebellar activity in PD, which has been suggested to be pathological or compensatory (Mirdamadi, 2016; Wu & Hallett, 2013). The higher range of the most distinctive frequency bands, on the other hand, might reflect impaired proprioceptive processing, which is also commonly associated with PD (Abbruzzese & Berardelli, 2003; Benatru et al., 2008; Jacobs & Horak, 2006). Noteworthy, our wavelet-based frequency spectrograms were only calculated up to 5 Hz. Since the largest gradients in some GradCAMs seem to exceed this range (Fig. 2), future investigation inspecting higher frequencies might show stronger correlations also with those frequencies. The frequency bands revealed by our GradCAMs have a slight overlap with those of the slow resting-tremor commonly associated with PD, which manifests between 4 – 7 Hz (Rivlin-Etzion et al., 2006). However, only two of our five best-performing models (#79, #183) had their largest gradients within this frequency range. Moreover, we checked for tremor frequencies in the hand motion of all participants. This revealed that two of the individuals with PD exhibited a tremor, one of them in only one hand. In both cases, the tremor frequency was around 5 Hz, as reported in the literature.

Thus, since only two out of 18 participants with PD exhibited a tremor, which in addition was located at the outer frequency limit of our spectrograms, we exclude tremor as potential cause for the high gradients in the individuals with PD. This suggests the selectivity for those frequency bands to stem from postural motion. Another trait that is shared between all GradCAMs constitutes the weight consistency across time. On average, there seems to be no relevant temporal information in the spectrograms for the network’s decision process. On the physiological side, this means that the frequency content of postural sway during quiet stance seems to remain stable over time, for individuals with PD and healthy adults alike. In terms of classification, in our case, this means that simpler classifiers trained on 1-D frequency data might be sufficient, with the additional advantage of even larger sample sizes, since singular time points could be used.

Our study was mostly limited due to the small number of participants. Even though the heterogeneity in our group of people with PD regarding their age, disease progression and dose of medication reflected the general population with early to mid-stage PD, our small sample size might have introduced biases. For instance, as can be seen in Table 1, even when based on medio-lateral COM data, classification performance of our models occasionally dropped below 70 %. This indicates that there were participants who shared characteristics across groups. However, given our generally robust results and the consistent imagery obtained from the GradCAMs, this shows that our methods were able to capture common traits that were unique to people with PD. If these results can be confirmed in large-scale studies, our deep learning model has the potential to reliably detect postural impairments in PD in the general population. Another limitation constitutes that all subjects with PD who participated in our study have already been diagnosed for a longer time and were already receiving treatment. If our model was able to find differences in people with de-novo PD, this would mark a large step towards assisting clinicians in initial assessments. In addition, distinguishability between the groups might be enhanced by having participants close their eyes during the recording or standing on non-firm ground, since due to their reduced proprioception, individuals with PD have been found to have increased reliance on vision (Abbruzzese & Berardelli, 2003; Benatru et al., 2008; Keijsers et al., 2005). Large-cohort studies with recently diagnosed individuals under more challenging conditions are currently in planning.

Lastly, there is evidence that dynamic balance control, e.g., in response to external stimuli, is better suited for distinction of people with PD from healthy controls (Benatru et al., 2008; Nardone & Schieppati, 2006). However, investigating dynamic balance scenarios usually requires much higher technical and experimental effort. Our models were not better at distinguishing between the groups based on data taken from the dynamic conditions of our previous studies (Engel, Student, et al., 2021; Student et al., 2022). This is remarkable, since it renders the bulk of the setup obsolete. Obtaining the trajectory of the COM during quiet standing only requires the Kinect, a single, cost-effective device. This means, our deep learning approach is able to classify individuals with PD from healthy controls with significantly reduced experimental effort when compared to conventional methods, requiring only 30 s or less of data recording.

### Conclusion

Using a convolutional neural network to classify individuals with early to mid-stage PD based on the frequency content of their body sway during quiet standing allowed us not only to predict postural instability in PD with excellent accuracy, but also revealed that postural impairments are reflected in specific frequency bands. Since these findings are backed by the literature, this proves our explainable AI approach (GradCAM) to provide meaningful insight into posturographic data. As our results can be achieved with short recording times and minimal experimental effort, this study design can easily and conveniently be applied on large scales. Overcoming current limitations, our method thus bears large potential to deepen our understanding of postural instability in PD and to facilitate clinical evaluation of the disease.

## Methods

### Participants

We assessed a group of 18 individuals with PD (age: range: [42-76]; mean ± standard deviation: 58.10 ± 8.66) in early to moderate disease stages (Hoehn and Yahr: ([1-3]; 1.94 ± 0.70 (Hoehn & Yahr, 1967) with a mean disease duration of 4.8 years ([0-15]; 4.79 ± 4.71)) who were diagnosed according to the Movement Disorder Society diagnostic criteria (Postuma et al., 2015). To be included into the study, individuals with PD had to be able to walk without any assistance and not have more than one reported fall in the year prior to the study. Also, no *Freezing of Gait* was to occur neither in the preliminary clinical examinations nor during the measurement. All individuals with PD were measured while being “on” their regular dose of dopaminergic medication (Levodopa Equivalent Daily Dose (LEDD): [105-1980]; 651.63 ± 529.97). Detailed information on the individuals recruited for our PD group can be found in our previous work (Student et al., 2022). For the control group (healthy controls, HC), we recruited fifteen age-matched healthy adults (age: [49-70]; 59.80 ± 6.45). General exclusion criteria were any existing neurological (e.g., neuropathies, epilepsy, multiple sclerosis, schizophrenia, severe, depression, dementia etc.) disorders or orthopedic conditions that might affect upright stance and balance control (e.g., hip, spine, knee, etc.). Potential cognitive impairment was evaluated prior to the study based on the Montreal Cognitive Assessment with a cut-off score of 24 point (Ciesielska et al., 2016). All subjects had normal or corrected to normal vision. All participants gave written informed consent prior to the experiment, including about the storage and processing of their data. Experimental procedures involving healthy individuals were approved by the Ethics Committee of the Psychology Department, University of Marburg. Research including individuals with PD was approved by the Ethics Committee of the Faculty of Medicine, University of Marburg (Case 77/19). All research was conducted in accordance with the Declaration of Helsinki.

### Experimental Setup

Experimental data for this study was acquired as part of two previous studies (Engel, Student, et al., 2021; Student et al., 2022). Participants stood on a Wii Balance board (WiiBB, Nintendo, Kyoto, Japan) to track their COP. Wearing no shoes, they were instructed to position their feet about shoulder width apart, about parallel on the ground. In one experimental condition, participants were to stand quietly with eyes open in a virtual 3-D environment, which consisted of a tunnel stretching in the anterior-posterior direction. During trials, they were instructed to stand relaxed with their arms hanging loosely at their sides. They had to fixate a target in the center of the far end of the tunnel to ensure their gaze remained straight ahead. In this manner, this condition simulated quiet standing in a natural environment with eyes open. One trial of measurement lasted for 30 seconds and was preceded and followed by resting periods where participants could relax and stretch as long as they needed. Each participant performed a total of 10 trials. To track their body motion, we used a Kinect v2 video-based motion tracking system (Microsoft, Redmond, WA, USA) which recorded the 3D-positions of 25 different ‘body joints’ as determined by an internal algorithm. The camera was located 210 cm in front of the participants and fixed at a height of 140 cm. The visual environment was presented through a head-mounted virtual reality headset (HTC Vive, HTC, New Taipei City, Taiwan). The frame rate was 90 Hz. The field of view extended over 110° in the vertical as well as in horizontal direction. Participants were secured by a harness which was attached to the ceiling. We ensured that the harness guaranteed subjects’ safety but was not providing lift during trials. For a more detailed description and depictions of the technical setup and experimental protocol, please refer to our previous work (Engel, Student, et al., 2021; Student et al., 2022).

### Data processing, architecture, and evaluation

Out of the WiiBB sensor data, we calculated the anterior-posterior (a-p) and medio-lateral (m-l) COP trajectories for each trial. The respective trajectories of the center of mass (COM) from the Kinect data were obtained based on the interpolated center positions of relevant body segments along with their attributed mass contributions, which were taken from anthropometric tables (Winter, 2009). Hence, our spatiotemporal data set consisted of two postural parameters, the COP and COM, with two spatial directions per parameter, the a-p and m-l directions of body sway, respectively. Data of each parameter and direction was resampled to 50 Hz using a custom-written Gaussian moving average filter with a symmetric window (sigma = 1/60 s), resulting in 1500 time points per 30 s-trial. We then performed a wavelet decomposition (generalized Morse wavelets, gamma: 3; 10 voices per octave; frequency range: [0.11 Hz - 5 Hz]) on all trajectory data to acquire time-frequency spectrograms for each trial. Subsequently, to obtain more data samples, the spectrograms were cut into 5 s-segments (250 time points), resulting in a total of 60 wavelet-based time-frequency spectrograms per subject (composed of 10 trials * 6 time segments) for each parameter and direction. Each spectrogram contained 2-D data (250 time points * 56 frequency bands) with one energy value associated with each time-frequency point, representing the respective frequency power. The resulting spectrograms thus resembled 56×250 px grayscale images with the energy values represented as pixel values (Fig. 3).

**Fig. 3.**
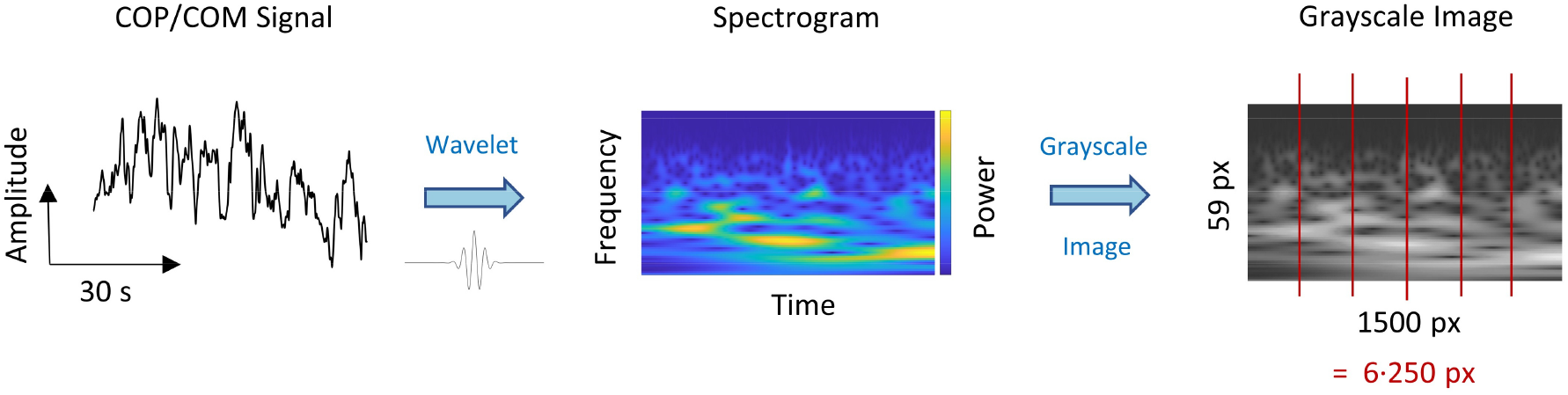
Data preprocessing. The time-course signals of the COP and COM trajectories underwent wavelet decomposition, resulting in spectrogram images with frequency power as a function of time and frequency. These spectrograms were then treated as grayscale images. Subsequently, each image was cut into six sections. The final samples constituted grayscale images with a size of 59×250 px.

Network training using the previously acquired spectrogram images was performed for each parameter (COP, COM) and direction (a-l, m-l) separately. Since one of our aims was to establish a system that can classify new subjects, the spectrograms from each group were split into train and validation sets on a per-subject basis. Hence, all 60 spectrogram images of each participant were assigned either entirely to the train or validation set, respectively. Due to technical issues, 10% of the total trials in the PD group and 3% of the total trials in the HC group needed to be discarded, resulting in some participants providing less than 60 samples (PD: n=7, HC: n=3). To avoid biases due to unequal group sizes, for each model we trained, only 15 out of the 18 subjects with PD were randomly selected, resulting in 15 subjects in both groups. Subject data was labeled binarily regarding group. Subsequently, 11 subjects from each group were assigned to the training set, while the remaining 4 subjects were assigned to the validation set (Fig. 4). Once assigned to the train and test sets, the pixel values of the spectrograms were normalized according to mean and standard deviation across all subjects. Importantly, the normalization coefficients obtained from the training set were applied to both train and test set. For cross-validation purposes and to facilitate generalizability, the per-subject assignment to each set was randomized before a new model was trained, resulting in different subjects in the respective train and test sets of each model we trained.

**Fig. 4.**
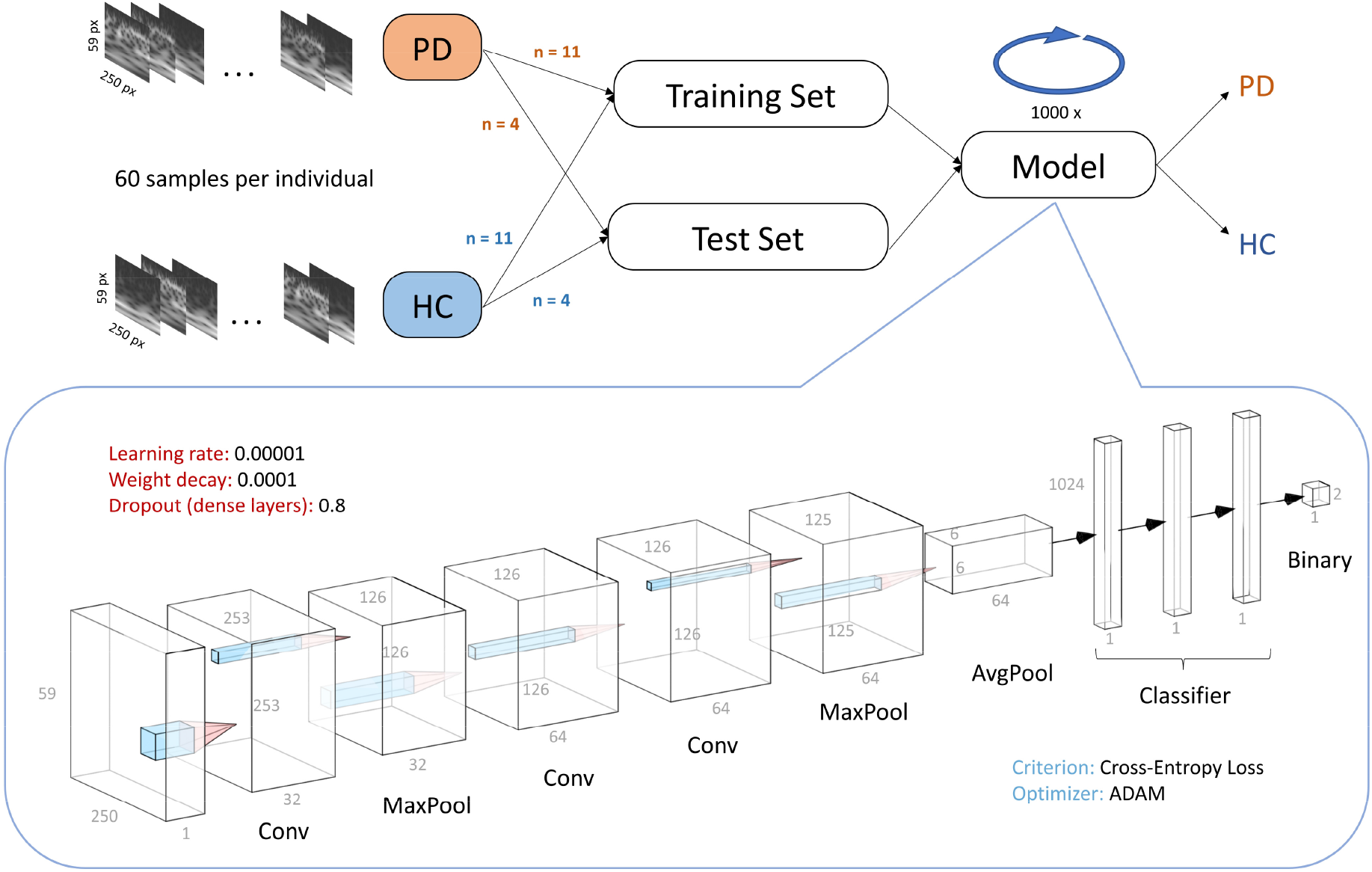
Data pipeline and architecture. Data was split into train and test sets on a randomized per-subject basis. For each model that we trained, data from 11 participants out of each group, consisting of the corresponding 60 sample images per participant, went into the training set. The remaining data went into the test set. The model was then trained and evaluated for 1000 epochs. We built a CNN with three convolutional and three pooling layers, feeding into a classifier network consisting of three fully-connected layers.

We built a custom CNN using the PyTorch framework (Paszke et al., 2019). The feature extractor network consisted of three convolutional and three pooling layers, the classifier consisted of three dense layers ending in a binary classification between PD and HC (Fig. 4). Each model was trained on full data batches over 1000 epochs with a learning rate of 0.0001 and slight regularization (weight decay = 0.001). We used an ADAM optimizer with cross-entropy-loss. For each parameter and direction, we trained a total of 250 models, each time with newly assigned subjects in the test and validation set. We then evaluated the models on peak and average performance for each parameter and direction. Evaluation criteria were predictive accuracy on test set data as well as corresponding receiver operating characteristics (ROC) curves with area under the curve (AUC) as performance measure. We also created a baseline performance to check whether the actual network performance was based on a true difference in the spectrograms between the groups. For this purpose, we also trained 250 models with randomly assigned labels and performed the same evaluation as with the correctly labeled data sets. Network performance was compared with the baseline performance using t-tests on the test set accuracy after 1000 epochs of training across all 250 models, respectively. We considered a p-value < 0.05 to reject the hypothesis that model performances between the shuffled and actual data sets came from the same distribution. To gain insight into the decision process of the trained models, we employed an explainable AI approach utilizing Gradient-weighted Class Activation Mapping (GradCAM, Selvaraju et al., 2020). GradCAMs identify the gradient information flow into the decision layer for the decoded class from each pixel of the input. The output can be visualized by heatmaps of the same size as the input image. These heatmaps indicate areas on which the model is focusing for classification, identifying which parts of the input image contribute the most to the class decision in the decision layer.

## Data Availability

All data and code produced in the present study are available upon request to the authors. They will be made available online soon.

## Acknowledgements

This research was funded by Deutsche Forschungsgemeinschaft: IRTG-1901, CRC/TRR-135 (Project No. 222641018), European Union: PLATYPUS, and Hessisches Ministerium für Wissenschaft und Kunst: The Adaptive Mind (TAM). A.P. is supported by a Marie Skłodowska-Curie Actions – H2020-MSCA-IF-2018 grant (ID: 832518, Project: MOVES).

